# Risk of Incident Cytopenia in Clonal Hematopoiesis

**DOI:** 10.1101/2024.09.30.24314665

**Authors:** James Brogan, Ashwin Kishtagari, Robert W. Corty, Yash Pershad, Caitlyn Vlasschaert, Brian Sharber, J. Brett Heimlich, Leo Luo, Yaomin Xu, Alexander G. Bick

## Abstract

Clonal hematopoiesis of indeterminate potential (CHIP) is an asymptomatic condition associated with elevated risk for myeloid neoplasms (MN). Patients with CHIP and cytopenia are at greater risk of MN. Quantifying the incidence of cytopenia and identifying risk factors among CHIP patients is critical for improving clinical management. We analyzed sequencing data from 805,249 participants in the NIH All of Us Research Program (AoU), Vanderbilt’s BioVU repository, and UK Biobank (UKB). Genetic mutations, laboratory values, and MN diagnoses were included in survival analyses to determine predictors of cytopenia in individuals with CHIP and matched controls. The cohort contained 9,374 CHIP cases and 24,749 controls. Cytopenia occurred in 13.5% of cases and 11.6% of controls (HR = 1.17, 95% confidence interval:

1.10 – 1.25, P=2.5 × 10^−6^). Cytopenia risk factors included smoking, male sex, variant allele frequency ≥ 0.20, age ≥ 65, mean corpuscular volume ≥ 100 femtoliters, red cell distribution width ≥ 15%, mutations in high-risk CHIP genes, and ≥ 2 CHIP mutations. In BioVU, 45% of participants with ≥ 4 risk factors progressed to cytopenia within two years. Individuals with CHIP and cytopenia progressed to MN at a rate of ~2% per year, compared to <0.1% per year for those without cytopenia. Longitudinal analysis across three cohorts demonstrated an increased risk of cytopenia in CHIP patients and identified those at highest risk. These findings suggest that cytopenia is a critical step in progression from CHIP to MN, underscoring its utility as an endpoint in cancer prevention trials for CHIP patients.

## Introduction

Clonal hematopoiesis (CH) is common among older individuals and is a risk factor for lymphoid and myeloid neoplasms (MN) as well as multiple other diseases of aging^1–7^. CH of indeterminate potential (CHIP) is a type of CH defined by somatic mutations in leukemia driver genes at a variant allele fraction (VAF) ≥ 2% in the absence of a diagnosed blood disorder or cytopenia^8^. Clonal cytopenia of undetermined significance (CCUS) differs from CHIP in that individuals with CCUS have a persistent unexplained cytopenia in the absence of MN. CHIP and CCUS are observed in 10% to 20% of individuals aged over 70 years of age^1–3,9^. Both CHIP and CCUS are premalignant lesions that confer increased risk of progression to MN. Previous estimates of MN progression have lumped CHIP and CCUS together, with a commonly quoted progression rate of ~0.5% per year^10,11^. However, emerging data suggests that these entities may confer distinct risk profiles. For example, cross-sectional analyses of individuals with CHIP and evidence of cytopenia based on a single blood draw have found that cytopenia confers increased risk of MN compared to individuals with CHIP who have normal blood counts^12–14^. Notably prior analyses did not employ longitudinal complete blood count data to differentiate between the two disease states. Therefore, the rate of progression from CHIP directly to MN may be lower than currently estimated as individuals may progress from CHIP to CCUS before developing MN.

Our ability to predict which individuals with CHIP will develop cytopenia is limited. Quantifying the incidence of cytopenia in individuals with CHIP and identifying features associated with progression to cytopenia can help improve current risk stratification models and identify individuals with CHIP who may benefit from closer monitoring or intervention^15^.

Demonstrating that cytopenia is in the causal pathway of CHIP progression to MN using longitudinal data is also extremely important for cancer prevention clinical trial design. Given the very low rate of progression from CHIP or CCUS to MN, extraordinarily large trial sample sizes would be required to demonstrate clinical benefit. However, cytopenia is much more common than MN. A cancer prevention trial in patients with CHIP seeking to test whether an intervention prevents the development of cytopenia would require a smaller sample size and be considerably more feasible to execute.

Here, we analyzed longitudinal complete blood count data from three large biobanks to quantify the incidence of cytopenia among individuals with CHIP. We identify genetic, demographic and laboratory values associated with increased risk of cytopenia in individuals with CHIP. We suggest that developing a cytopenia is a required step in the progression from CHIP to MN, highlighting its clinical utility as a clinical trial endpoint.

## Methods

### Cohort descriptions

Participant data was obtained from three large observational cohorts: the All of Us Research Program (AoU), UK Biobank (UKB) and Vanderbilt’s BioVU biorepository. AoU is an ongoing US-based observational cohort study^16^. AoU whole genome sequencing (WGS) data was available for 243,609 participants who were enrolled from 2017 to 2022. These participants have linked health outcome data from participant survey questionnaires and electronic health records harmonized to the Observational Medical Outcomes Partnership (OMOP) Common Data Model (CDM) at enrollment and follow-up time points. WGS was performed using Illumina PCR-free whole genome technology and sequenced on the NovaSeq platform to a median sequencing depth of 40x. The median age at enrollment for this cohort is 53 years old (interquartile range, 37 to 65).

BioVU is Vanderbilt’s biorepository of DNA extracted from discarded blood collected during routine clinical testing and linked to de-identified medical records derived from Vanderbilt’s electronic medical record^17^. WGS data was available for 107,607 adult participants enrolled from 2006 to 2023 who have linked electronic health record data harmonized to the OMOP CDM at enrollment and each subsequent healthcare encounter. WGS was performed using Illumina PCR-free whole genome sequencing technology and sequenced on the NovaSeq platform to a median sequencing depth of 35x. The median age at enrollment for this cohort is 50 years old (interquartile range, 34 to 62).

UKB is a UK-based observational cohort study. UKB whole exome sequencing (WES) was available for 454,033 participants aged 40 to 70 at time of DNA collection^18^. Participants were enrolled from 2007 to 2010 and have questionnaire, physical measurement, laboratory, and medical imaging data available at enrollment and follow-up time points^19^. Health outcomes since enrollment are tracked from hospitalization general practice health records and death and cancer registries. WES was performed to a median sequencing depth of 40x across sites^20^. The median age at enrollment for this cohort is 58 years old (interquartile range, 50 to 63). The characteristics of eligible study participants included in our study from each cohort are shown in Table S1 to S3.

### Study design

We conducted a case-control cohort study of participants from AoU, BioVU, and UKB. Individuals were eligible for the study if they had sequencing and multi-timepoint complete blood count (CBC) data, without evidence of cytopenia, acute myeloid leukemia (AML), myelodysplastic syndrome (MDS), or myelofibrosis prior to sequencing. Multi-timepoint CBC was defined as at least three CBC measurements, including one within a year of sequencing and two on or after the date of sequencing. The final CBC measurement had to occur at least 120 days after sequencing or the first CBC measurement, whichever came later (Figure S1). CBC measurements occurring greater than one year before sequencing were not included in analysis. Participants with CHIP were matched 1:3 with controls on age, sex, and smoking status. The follow-up period commenced at the date of sequencing and terminated at the earliest occurrence of persistent cytopenia, myelofibrosis, MDS, AML, or last CBC. The primary outcome of interest was incident cytopenia any time after study enrollment.

### Calling CHIP

Somatic mutations in 58 canonical CHIP driver genes were identified from read-level WGS data in AoU and BioVU, and WES data in UKB, using Genome Analysis Toolkit Mutect2^21^ and ANNOVAR^22^ as previously described^11^.

### CHIP, cytopenia, and MN definitions

CH was defined by the presence of somatic mutations in a canonical CHIP driver gene at a VAF ≥ 2% in AoU, BioVU, and UKB. CH without cytopenia or a diagnosed blood disorder was classified as CHIP. Participants with CHIP and an incident cytopenia were not classified as CCUS because bone marrow biopsy reports were not available for all participants. Cytopenias were defined by using a modified version of World Health Organization criteria^8^ (anemia: hemoglobin < 12.0 g/dL (females) or 13.0 g/dL (males); thrombocytopenia: platelets < 150,000 cells/μL; and leukopenia: white blood cell count < 3,700 cells/μL). Cytopenias were only deemed to be persistent if there were two consecutive observations of a cytopenia in a single lineage at least 120 days apart without an intervening normal measurement (Figure S1). The date of cytopenia was the first occurrence of the cytopenia that persisted for at least 120 days. MN was defined as AML, MDS, or myelofibrosis. Essential thrombocythemia and polycythemia vera were excluded from the definition of MN in this study because of their phenotypic heterogeneity and the high likelihood of misclassification in electronic health record data^23^.

### Data curation

Date of birth and death, sex, race, laboratory values, self-reported smoking history, and International Classification of Diseases, Ninth and Tenth Revision (ICD-9, ICD-10), codes were extracted across all three biobanks. All laboratory measurement variables were harmonized to a common unit of measure and screened for outlier values. ICD-9 and ICD-10 codes are listed in Table S4.

### Statistical methods

Statistical analyses were performed using Python (v3.10.12) and survival analyses were performed using R statistical software. Figures were made with matplotlib (v3.7.2)^24^ and R (version 4.2.1). All statistical tests were two-sided with statistical significance determined by a P value < 0.05. Cumulative incidence of cytopenia and MN were estimated by using a competing risks approach^25^. The competing risk for incident cytopenia was MN and the competing risk for MN was death. Cox proportional-hazards models were used to estimate risk of incident cytopenia for demographic, genetic and laboratory features.

## Results

Sequencing data were available for 805,249 participants. 162,017 participants, including 9,375 with CHIP, met inclusion criteria (Figure 1). Amongst patients who did not meet inclusion criteria, 615,505 were dropped because of insufficient serial CBC data collected during routine care. The final matched case-control cohort contained 9,374 cases and 24,749 controls (Figure 1, Figure S2). Characteristics of cases and controls are detailed in Table 1. There was no difference in age, sex, or smoking history between cases and controls in the composite dataset or in each cohort. However, there were inter-cohort differences (Table S1 to S3). Most notably, the median age of cases in UKB was 62.4 years (interquartile range, 57.5 – 66.2) which was significantly younger than the median age of 69.2 years (interquartile range, 60.9 – 75.6) in AoU and 65.2 years (interquartile range, 55.6 – 73.0) in BioVU. Median time at risk was 5.08 years in cases (interquartile range, 2.51 to 6.50) and 5.16 years in controls (interquartile range 2.53 to 6.54). Incident cytopenia occurred in 1,269 (13.54%) cases and 2,882 (11.64%) controls, with anemia being the major cause of cytopenia. Fine-Gray modeling for incident cytopenia with competing risk of MN demonstrated that cases with CHIP had a significantly increased risk of incident cytopenia compared to matched controls (HR = 1.17, 95% confidence interval: [1.10 – 1.25], P=2.5 × 10^−6^) with consistency across all three cohorts (Figure 2A and 2B). This consistency of effect was particularly notable given the cumulative incidence of cytopenia over two years varied considerably across the cohorts, with 15.9% of cases in BioVU and 13.1% in AoU compared to just 2.9% of cases in UKB.

**Table 1.** Characteristics of cases and controls.^*^

| Characteristic | Cases<br>(N=9,374) | Controls<br>(N=24,749) |
| --- | --- | --- |
| Age – median [IQR], year | 63.6 [57.7, 68.4] | 63.1 [57.1, 67.7] |
| Female – no. (%) | 5,237 (55.9) | 13,898 (56.2) |
| Any smoking history – no. (%) | 4,812 (51.3) | 12,639 (51.1) |
| Laboratory values<sup>†</sup> |  |  |
| Hemoglobin – median [IQR], g/dL | 14.0 [13.2, 14.9] | 14.0 [13.2, 14.9] |
| Platelet count – median [IQR], (10<sup>9</sup> cells/L) | 247 [210, 290] | 244 [208, 286] |
| White blood cells – median [IQR], (10<sup>9</sup> cells/L) | 6.9 [5.7, 8.3] | 6.7 [5.6, 8.1] |
| Mean corpuscular volume – median [IQR], fL | 91.0 [88.1, 94.0] | 91.0 [88.1, 94.0] |
| Red cell distribution width – median [IQR], % | 13.5 [13.0, 14.1] | 13.4 [12.9, 14.0] |
| Follow-up – median [IQR], year<sup>‡</sup> | 5.1 [2.5, 6.5] | 5.2 [2.5, 6.5] |
| Type of incident cytopenia<sup>§</sup> |  |  |
| Anemia – no. (%) | 1,013 (10.8) | 2,299 (9.3) |
| Thrombocytopenia – no. (%) | 260 (2.8) | 587 (2.4) |
| Leukopenia – no. (%) | 136 (1.5) | 271 (1.1) |
| Incident cytopenia – no. (%) | 1,269 (13.5) | 2,882 (11.6) |
| Incidence of cytopenia – per 1,000 person-years | 29 | 25 |
| Time to cytopenia – median [IQR], years | 2.3 [0.7, 4.2] | 1.9 [0.5, 3.0] |
| Incident AML, MDS, MF – no. (%) | 98 (1.1) | 92 (0.4) |
| AML – no. (%) | 39 (0.4) | 39 (0.2) |
| MDS – no. (%) | 47 (0.5) | 52 (0.2) |
| MF – no. (%) | 12 (0.1) | 1 (0.004) |
| Time from cytopenia to MN<sup>¶</sup> – median [IQR], years<sup>#</sup> | 1.5 [0.6, 3.2] | 2.6 [0.7, 5.5] |
| Death – no. (%) | 1,045 (11.2) | 2,204 (8.9) |

IQR: interquartile range; AML: acute myeloid leukemia; MDS: myelodysplastic syndrome; MF: myelofibrosis; MN: myeloid neoplasm.
\* Cases and controls were matched 3:1 on age ± 3 years, sex, any smoking history.
<sup>†</sup> Hematologic measurements were obtained from complete blood count obtained nearest to time of sequencing.
<sup>‡</sup> Follow-up time is the number of years from sequencing to death or last follow-up in each cohort, whichever is earliest.
<sup>§</sup> Cytopenia definitions: anemia (hemoglobin < 12.0 g/dL for females or 13.0 g/dL for males), thrombocytopenia (platelet count < 150,000 cells/ μL), leukopenia (white blood cell count < 3,700 cells/ μL). Cytopenias were only counted if there were two consecutive observations of a cytopenia in a single lineage at least 120 days apart without an intervening normal measurement.
<sup>||</sup> Times only reported for participants who developed cytopenia without subsequent myeloid neoplasm or cytopenia before myeloid neoplasm.
<sup>¶</sup> Myeloid neoplasms are defined as acute myeloid leukemia, myelodysplastic syndrome, myelofibrosis.
<sup>#</sup> Times reported for participants who developed cytopenia prior to myeloid neoplasm.

**Figure 1.**
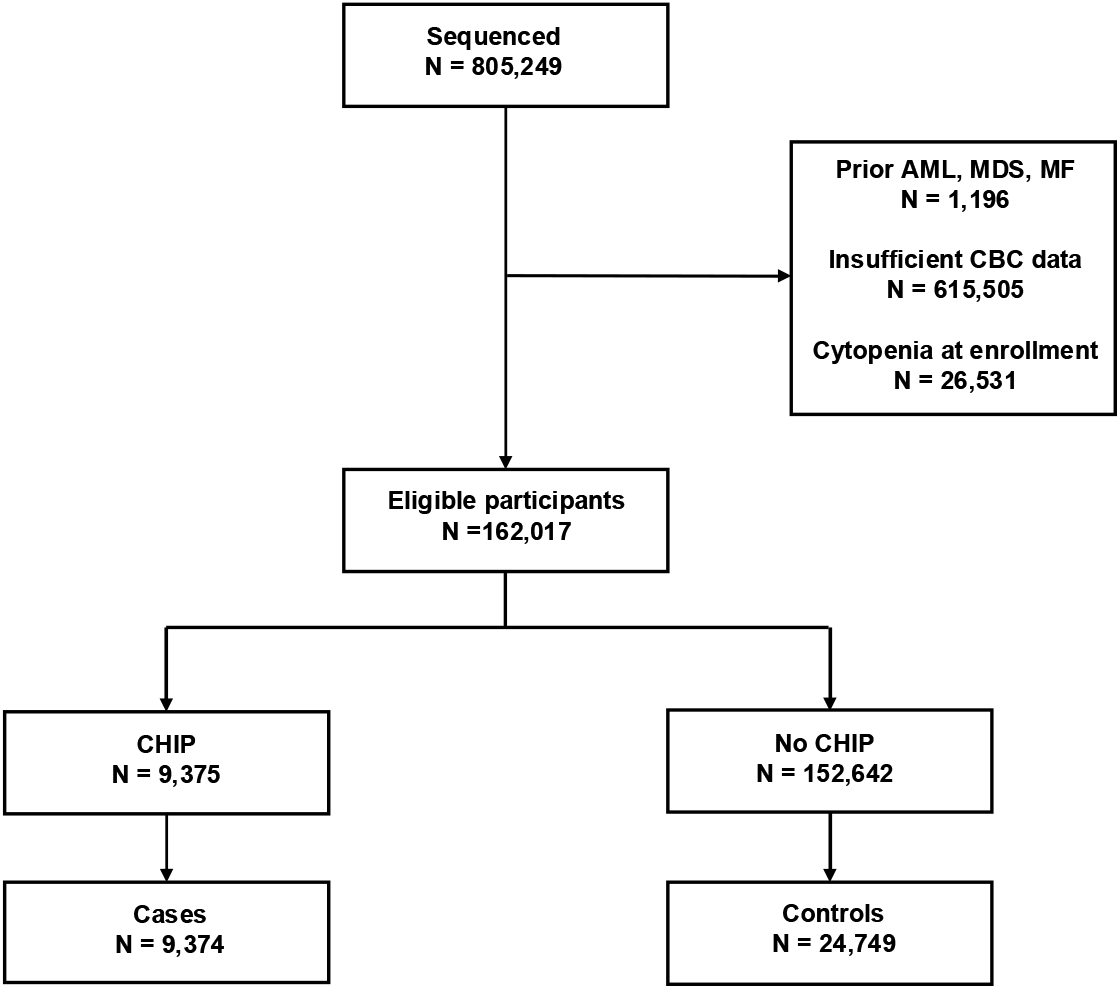
Flow diagram showing selection of cases and controls. Abbreviations: AML = acute myeloid leukemia; MDS = myelodysplastic syndrome; MF = myelofibrosis; CBC = complete blood count; CHIP = clonal hematopoiesis of indeterminate potential. Participants were screened from the All of Us Research Program (N=243,609), Vanderbilt’s BioVU biorepository (N=107,607), and UK Biobank (N=454,033). Participants were excluded for prior AML, MDS or MF diagnoses, insufficient CBC data, or cytopenia at enrollment. Cases and controls were matched 1:3 on age ± 3 years, gender, and any history of smoking within their respective cohort. The same control was able to be matched to multiple cases.

**Figure 2.**
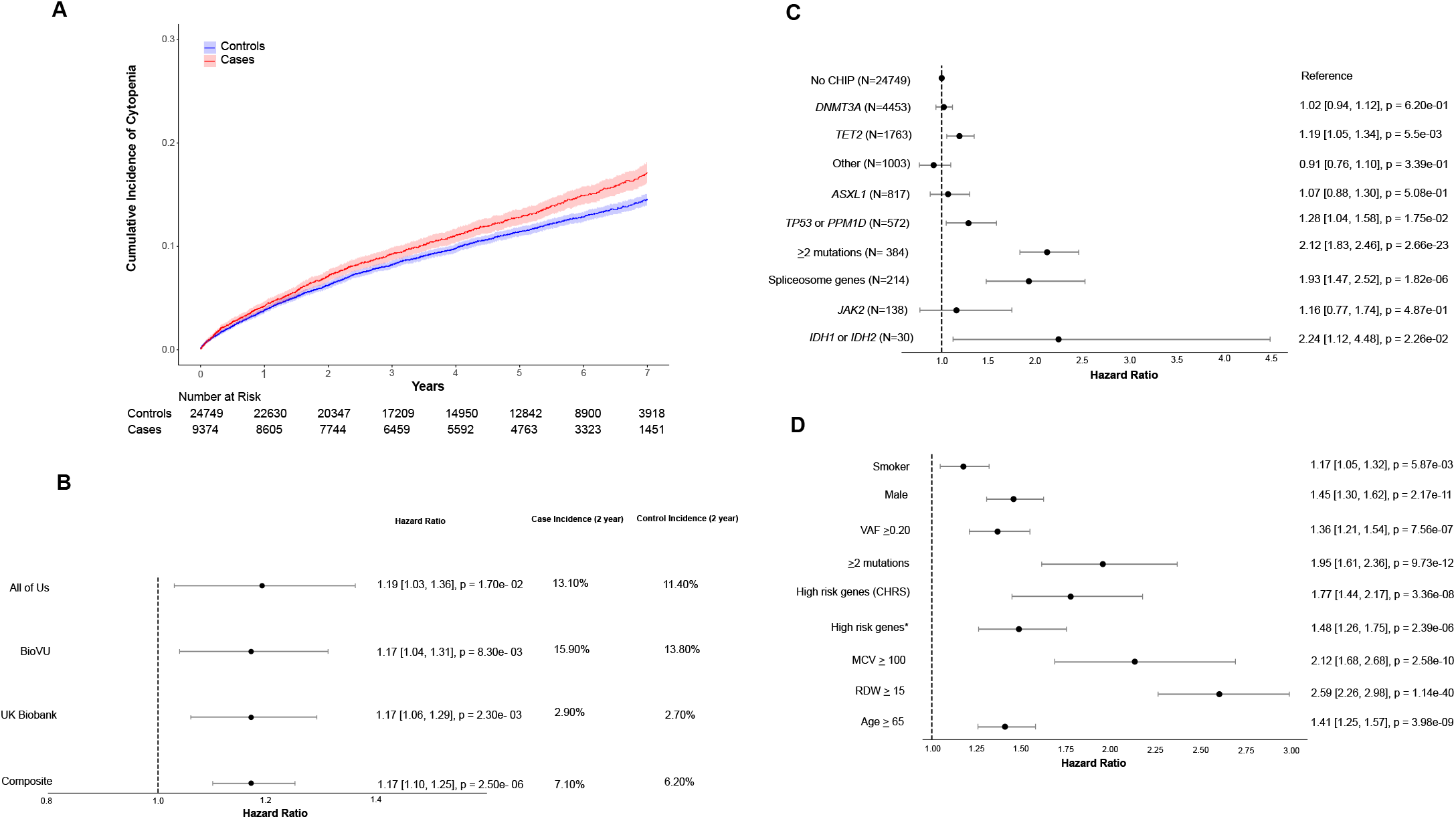
Features influencing risk of cytopenia in participants with CHIP. Abbreviations: VAF = variant allele fraction; CHRS = clonal hematopoiesis risk score; MCV = mean corpuscular volume (femtoliters); RDW = red cell distribution width (%); CHIP = clonal hematopoiesis of indeterminate potential. (A) Cumulative incidence of cytopenia in participants with CHIP compared to matched controls across the All of Us Research Program, Vanderbilt’s BioVU biorepository, and UK Biobank. Controls were matched for age ± 3 years, gender and any history of smoking. (B). Risk of incident cytopenia in participants with clonal hematopoiesis of indeterminate potential (CHIP) compared to matched controls across the All of Us Research Program, Vanderbilt’s BioVU biorepository, UK Biobank, and all three cohorts combined with their respective cumulative incidence of cytopenia two years after enrollment. Forest plot indicates HR and 95% confidence intervals. (C) Univariate Cox regression analyses for incident cytopenia by specific CHIP genotypes at time of enrollment for participants with CHIP and controls without CHIP serving as the reference group. (D) Univariate Cox regression analyses for incident cytopenia by baseline characteristic at time of enrollment for participants with CHIP without adjustment. The variable high-risk genes (CHRS) indicates a participant had at least one mutation in the following genes: *SRSF2, SF3B1, ZRSR2, IDH1, IDH2, FLT3, RUNX1*, or *JAK2*. The variable high-risk genes^*^ indicates a participant had at least one mutation in the following genes: *TP53, PPM1D, SF3B1, SRSF2, U2AF1, ZRSR2, IDH1*, or *IDH2*.

Cox regression analyses controlling for cohort of enrollment were conducted to identify which CHIP driver mutations conferred the highest risk of incident cytopenia. CHIP driven by a mutation in *TET2* was found to confer an increased risk of incident cytopenia compared to individuals without CHIP (HR = 1.19, 95% confidence interval: [1.05 – 1.34], P=5.6 × 10^−3^), while CHIP driven by *DNMT3A, ASXL1*, and *JAK2* did not (Figure 2C). Aggregating CHIP driver mutations by mechanism demonstrated increased risk of incident cytopenia among participants with driver mutations in the *TP53* pathway (*TP53* or *PPM1D*; HR = 1.28, 95% confidence interval: [1.04 – 1.58], P=1.8 × 10^−2^), in spliceosome genes (*SF3B1, SRSF2, U2AF1*, or *ZRSR2*; HR = 1.93, 95% confidence interval: [1.47 – 2.52], P=1.8 × 10^−6^), and CHIP driven by a mutation in AML-like genes (*IDH1* or *IDH2*; HR = 2.24, 95% confidence interval: [1.12 – 4.48], P=2.3 × 10^−2^).Individuals with two or more CHIP mutations were also at significantly increased risk for incident cytopenia (HR = 2.12, 95% confidence interval: [1.83 – 2.46], P=2.7 × 10^−23^). All effects were directionally consistent across cohorts (Figure S3).

Factors that increase risk for incident cytopenia were assessed using univariate Cox regression analysis controlling for cohort of enrollment. Smoking status, male sex, VAF ≥ 0.20, ≥ 2 CHIP mutations, ≥ 1 high-risk mutation^14^ (*SRSF2, SF3B1, ZRSR2, IDH1, IDH2, FLT3, RUNX1, JAK2*), mean corpuscular volume (MCV) ≥ 100 femtoliters, red cell distribution width (RDW) ≥ 15%, and age ≥ 65 years all demonstrated statistical significance (Figure 2D). These factors were directionally consistent across all three cohorts (Figure S4). In AoU, male sex, MCV ≥ 100 femtoliters, and RDW ≥ 15% conferred increased risk of cytopenia. In BioVU and UKB, all identified risk factors for incident cytopenia demonstrated significance except for VAF in BioVU and smoking status in UKB.

High risk features were defined as age ≥ 65 years, male gender, ≥ 2 CHIP mutations, MCV ≥ 100 femtoliters, RDW ≥ 15%, or the presence of any high-risk CHIP mutations. Individuals with multiple high-risk features had a significantly greater risk of incident cytopenia (Figure 3A and 3B). This increase in risk of incident cytopenia among participants with multiple high-risk features was consistent across cohorts (Figure S5). For example, BioVU participants with CHIP and no high-risk features had an 8.1% incidence of cytopenia at 2 years (95% confidence interval, 5.6% – 11.8%). This increased to 21.0% among participants with 2 or 3 risk factors (95% confidence interval, 18.0% – 24.4%) and 45.1% among participants with 4 or 5 risk factors (95% confidence interval, 33.3% – 58.9%) as demonstrated in Figure 3C.

**Figure 3.**
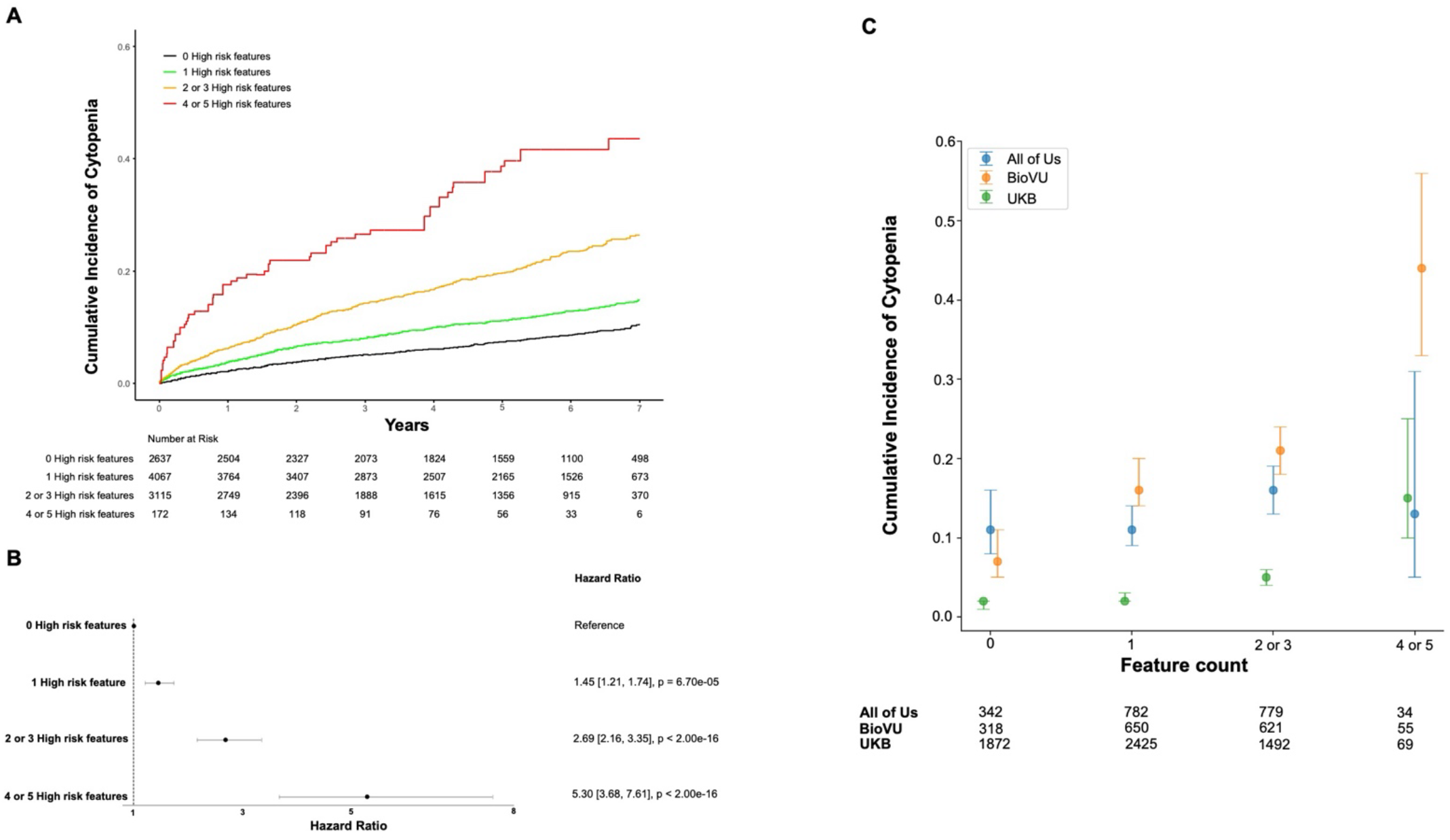
Risk of incident cytopenia stratified by participant risk factor profiles. (A) Cumulative incidence curve for cytopenia in participants with clonal hematopoiesis of indeterminate potential (CHIP) across All of Us Research Program, Vanderbilt’s BioVU biorepository, and UK Biobank stratified by the number of high-risk features they had at time of enrollment. High risk features were defined as age ≥ 65 years, male gender, ≥ 2 CHIP mutations, mean corpuscular volume ≥ 100 femtoliters, red cell distribution width ≥ 15%, or the presence of at least one high-risk CHIP mutation (*SRSF2, SF3B1, ZRSR2, IDH1, IDH2, FLT3, RUNX1*, or *JAK2*). (B) Hazard ratios for incident cytopenia were calculated for high-risk feature strata using Cox proportional-hazards models adjusted for cohort, sex, gender, and smoking history. Hazard ratios were calculated in a model with zero high-risk features as the reference population. Forest plot indicates HR and 95% confidence intervals. (C) Cumulative incidence of cytopenia two years after enrollment in participants with CHIP in each biobank stratified by number of high-risk features.

Among 34,123 cases and controls, there were 190 (0.56%) incident cases of MN. Participants with incident cytopenia had significantly higher risk of subsequent MN compared to those without cytopenia among both cases (HR = 24.3, 95% confidence interval: [6.6 – 89.0], P=2.0 × 10^−16^) and controls (HR = 44.6, 95% confidence interval: [18.8 – 106], P=2.0 × 10^−16^), with the highest risk of developing a hematologic malignancy taking place in cases (Figure 4A and 4B). There were 8,105 cases who did not develop a cytopenia of which 33 (0.4%) developed a MN at a median of 4.9 years (interquartile range, 2.4 to 6.5). Among 1,269 cases who developed an incident cytopenia, 56 (4.4%) developed a MN at a median of 2.3 years (interquartile range, 0.7 to 4.2). Controls who developed a cytopenia had a significantly greater risk of developing a hematologic malignancy compared to cases who did not develop a cytopenia (HR = 8.3, 95% confidence interval: [2.2 – 30.4], P=2.0 × 10^−16^). In participants who developed a cytopenia before MN or did not progress to MN during follow-up, median time to cytopenia was 2.3 years (interquartile range, 0.7 – 4.2) in cases and 1.91 years (interquartile range, 0.5 – 3.0) in controls. Participants who developed a cytopenia and progressed to MN had a median time from cytopenia to MN of 1.5 years (interquartile range, 0.6 – 3.2) in cases and 2.6 years (interquartile range, 0.7 – 5.5) in controls.

**Figure 4.**
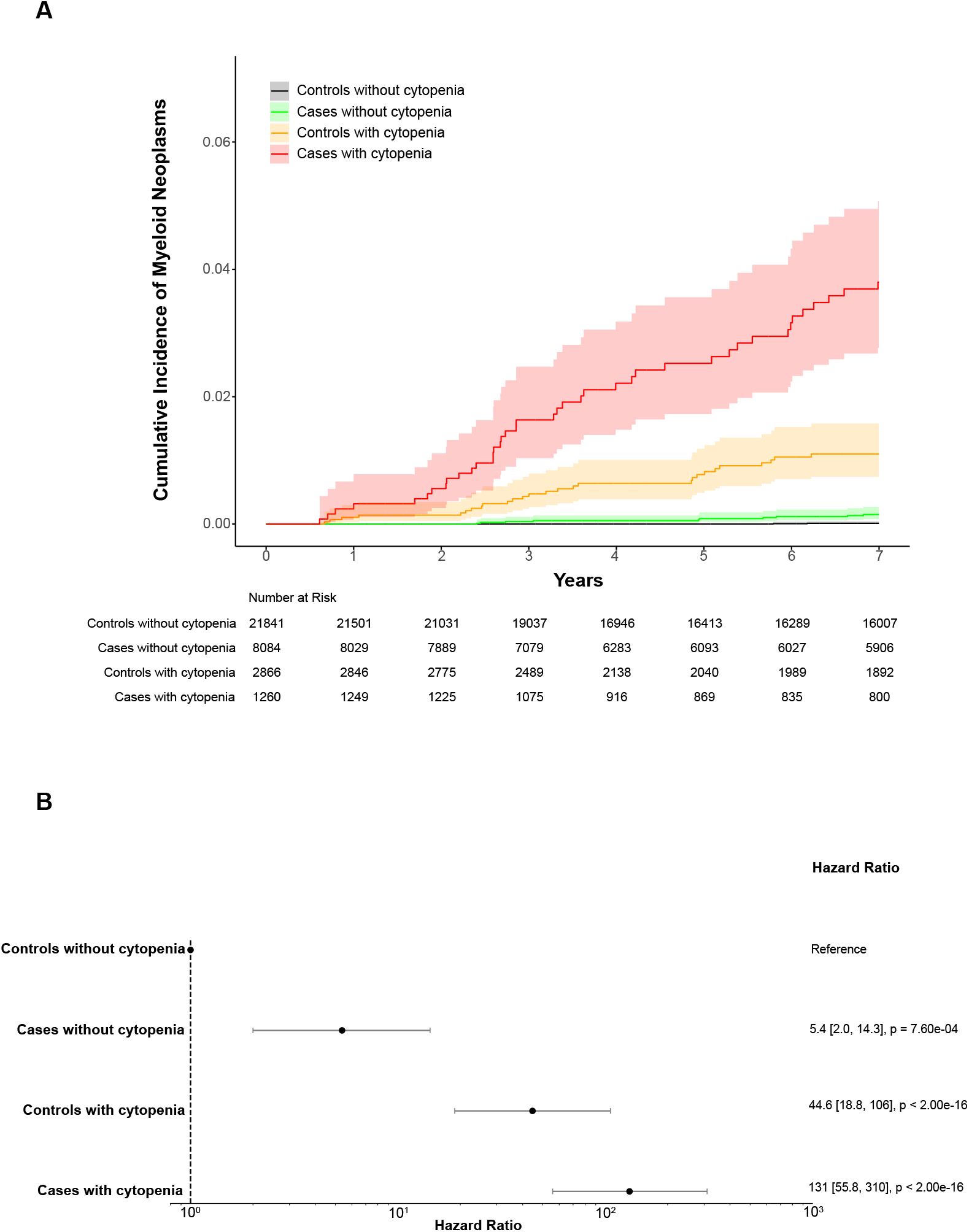
Risk of incident AML, MDS, MF stratified by CHIP status at enrollment and incident cytopenia. Abbreviations: AML = acute myeloid leukemia; MDS = myelodysplastic syndrome; MF = myelofibrosis; CHIP = clonal hematopoiesis of indeterminate potential. (A) Risk of incident AML, MDS, MF in participants with CHIP compared to matched controls across All of Us Research Program, Vanderbilt’s BioVU biorepository, and UK Biobank. (B) Hazard ratios for incident AML, MDS, MF were calculated for cases and controls stratified by incident cytopenia using Cox proportional-hazards models adjusted for cohort, sex, gender, and smoking history. Hazard ratios were calculated in a model with controls who did not develop an incident cytopenia as the reference population. Forest plot indicates HR and 95% confidence intervals.

## Discussion

Longitudinal analysis of 9,374 participants with CHIP across three distinct population-based cohorts demonstrated significantly higher incidence of persistent cytopenia in participants with CHIP than matched controls. Further analysis identified risk factors for developing persistent cytopenia in patients with CHIP and refined estimates of myeloid malignancy progression among those with and without a cytopenia and CHIP. These findings should be reassuring to both patients and clinicians caring for patients with CHIP as it highlights cytopenia as a biomarker in the causal pathway to malignancy. These observations permit several conclusions.

First, patients with CHIP have a substantial annual risk of developing cytopenias, ranging from 1-15% in the UK Biobank to 10-35% in the US-based All of Us and BioVU cohorts with degree of risk depending on the number of high-risk features. This increased risk of cytopenia among patients with CHIP has long been presumed, implicit in the conceptual framework of progression from CHIP to CCUS to MN but has not previously been quantified. The relative risk of cytopenia was remarkably consistent across all three cohorts. The consistency of effects across disparate cohorts provides confidence that the observed signal represents the underlying biological process rather than an idiosyncrasy of one cohort. Given the substantial risk of cytopenia, we would suggest that patients with multiple high-risk features are regularly monitored for cytopenia progression. Modeling the optimal timing for CBC monitoring is an important future direction.

Second, there were notable differences in the rate of cytopenia across the cohorts. Several factors may contribute to this difference. First, UK Biobank participants have been shown to be substantially healthier than the UK population in general^26^, while participants from a healthcare system such as those in BioVU were substantially less healthy than the US population in general. Second, UK Biobank participants who were eligible for this study were younger than those in the All of Us and BioVU cohorts. Third, the number of CBCs per participant differed by cohort. Different practice patterns in CBC collection may contribute to different rates of cytopenia detection across the cohorts.

Third, we found the absolute risk of developing a MN among participants with CHIP in the absence of cytopenia to be 0.05% – 0.1% per year, with the upper bound being ten-fold lower than the 0.03% – 1% annual risk previously reported^1,11^. Participants with CHIP who do not have cytopenia in our data have a risk of less than 0.1% per year of developing a MN, which is nearly identical to individuals without CHIP who do not develop a cytopenia. Furthermore, individuals with detectable CHIP mutations who went on to develop cytopenia and MN had a median latency period of approximately 5 years, consistent with latency periods before AML diagnosis in prior studies^5,27^. This latency period preceding cytopenia and MN could serve as an opportunity for monitoring and interventional studies.

Fourth, the existence of high-risk subsets of individuals with CHIP that can be identified based on a combination of routine clinical and genomic features enables the clinical development of therapies that prevent cytopenia progression. For example, to conduct a clinical trial for a therapeutic intervention that reduces the risk of progression from CHIP to cytopenia by a factor of 2, the study would require 868 patient-years of follow up time in a population similar to BioVU enrolling all CHIP participants with a 10% average annual risk of incident cytopenia. However, if the same study only enrolled CHIP participants with 4 or more risk factors, translating to a 38% average annual risk of incident cytopenia, only 174 patient-years of follow up would be required.

Lastly, prognostic variables used in clonal hematopoiesis risk score (CHRS) to calculate risk of progression from CHIP or CCUS to MN are highly concordant with the risk factors predicting CHIP to cytopenia^14^. High-risk mutations, ≥ 2 mutations, VAF ≥ 0.20, RDW ≥ 15%, MCV ≥ 100 femtoliters, and age ≥ 65 years demonstrated prognostic value in CHRS and were used as high-risk features in our analysis. Spliceosome genes including *SRSF2* and *SF3B1* have been shown to be associated with increased MCV and RDW as well as decreased platelet count and hemoglobin^13,28^, consistent with our results. The two prior studies both relied on the UK Biobank which was incorporated into our analyses as one of the three cohorts. Furthermore, splicing mutations and ≥ 2 mutations were found to be key adverse prognostic factors in progression of CCUS to MN^12,29–31^. In extending these findings to two additional US-based cohorts we reassuringly find that the results are applicable to US patient populations presenting for care in US health care settings.

Our study has several limitations. First, we derived CHIP calls based on both WES and WGS, this approach carries a lower depth of coverage compared to targeted sequencing approaches^32^. Our analysis reduced sensitivity for detecting CHIP clones that make up only a small fraction of the blood. Extending our analyses to small CHIP clones is an important area of future study. Second, our study did not include somatic copy number alterations, which are known to accumulate with age and are associated with subsequent development of blood count abnormalities and mortality from hematologic malignancy^33^. Future risk models could be improved by incorporating these genetic factors. Third, we identified a subset of patients who develop cytopenia without CHIP and subsequently develop MN. We posit that these could be driven by somatic copy number alterations or small CHIP clones which were undetected in this study. Addressing these limitations in future research will be crucial for a more comprehensive understanding of the relationship between CHIP, cytopenia and MN.

In conclusion, our data support the hypothesis that somatic mutations in CHIP driver genes increase risk of incident cytopenia. Our findings enhance existing risk models to enable longitudinal risk stratification and personalized clinical risk prediction among individuals with CHIP. We propose that cytopenia-free months could be a key endpoint for future cancer prevention clinical trials for patients with CHIP given that our data shows that cytopenia is a required step for malignancy progression. Our ability to identify individuals at the highest risk of developing cytopenia and subsequent MN will enable the efficient design and execution of such trials.

## Data Availability

All data produced in the present study are available upon reasonable request to the authors.

## Acknowledgements

This work was supported by grants (DP5OD029586, R01AG088657, R01AG083736, P30CA068485) from the National Institutes of Health (NIH), the Burroughs Wellcome Fund Career Award for Medical Scientists, the Edward P. Evans Foundation, the Pew Charitable Trusts and the Alexander and Margaret Stewart Trust, and a Hevolution/AFAR New Investigator Award in Aging Biology and Geroscience Research to AGB. JB was supported by the American Society of Hematology HONORS Award. This research was conducted using the UK Biobank Resource under Application Number 43397. The All of Us Research Program is supported by the National Institutes of Health, Office of the Director: Regional Medical Centers: 1 OT2 OD026549; 1 OT2 OD026554; 1 OT2 OD026557; 1 OT2 OD026556; 1 OT2 OD026550; 1 OT2 OD 026552; 1 OT2 OD026553; 1 OT2 OD026548; 1 OT2 OD026551; 1 OT2 OD026555; IAA #: AOD 16037; Federally Qualified Health Centers: HHSN 263201600085U; Data and Research Center: 5 U2C OD023196; Biobank: 1 U24 OD023121; The Participant Center: U24 OD023176; Participant Technology Systems Center: 1 U24 OD023163; Communications and Engagement: 3 OT2 OD023205; 3 OT2 OD023206; and Community Partners: 1 OT2 OD025277; 3 OT2 OD025315; 1 OT2 OD025337; 1 OT2 OD025276. BioVU is supported by NIH award UL1 TR002243 from the National Center for Advancing Translational Sciences. The BioVU Whole Genome Sequencing was supported by the Alliance for Genomic Discovery. We gratefully acknowledge All of Us, UK Biobank and BioVU participants for their contributions, without whom this research would not have been possible. The contents of this manuscript are solely the responsibility of the authors and do not necessarily represent official views of the NIH.

## Authorship

Contribution: JB, AK, RWC, YP and AGB conceived and designed the study. JB performed primary analysis and prepared figures and tables. JB, AK, RWC, YP and AGB contributed to interpretation of the data. BH and LL assisted with refinement of the study design. YX and AGB contributed to data acquisition. RWC, YP, CV and BS performed somatic mutation calls in the All of Us Research Program, UK Biobank, and Vanderbilt’s BioVU biorepository. JB, AK and AGB wrote the first draft of the manuscript which was critically revised for important intellectual content with feedback from all authors. Conflict-of-interest disclosure: AGB is on the scientific advisory board and has received personal fees from TenSixteen Bio unrelated to the present work.

Correspondence: Alexander G. Bick, Division of Genetic Medicine, Vanderbilt University Medical Center, 2200 Pierce Ave, Nashville, TN 37232.

## Notes

### Author Declarations

IRB of Vanderbilt University Medical Center waived ethical approval for this work

