## Supplement for "Risk of Incident Cytopenia in Clonal Hematopoiesis"

**Table of Contents**

I. **Supplemental Tables**

Table S1 – Characteristics of cases and controls in the All of Us Research Program

Table S2 – Characteristics of cases and controls in Vanderbilt’s BioVU biorepository

Table S3 – Characteristics of cases and controls in UK Biobank

Table S4 – ICD-9 and ICD 10 codes used to define myeloid neoplasms

II. **Supplemental Figures**

Figure S1 – Multi-timepoint complete blood count eligibility and cytopenia criteria

Figure S2 – Flow diagram showing selection of cases and controls across cohorts

Figure S3 – Risk of incident cytopenia by genotype across cohorts

Figure S4 – Risk of incident cytopenia by baseline characteristic across cohorts

Figure S5 – Risk of incident cytopenia stratified by participant risk factor profiles across cohorts

**Table S1. Characteristics of cases and controls in the All of Us Research Program.^*^**

| Characteristic | **Cases**  **(N=1,937)** | **Controls**  **(N=5,109)** |
| --- | --- | --- |
| Age – median [IQR], year | 69.2 [60.9, 75.6] | 68.2 [59.8, 74.7] |
| Female – no. (%) | 1,124 (58.0) | 2,983 (58.4) |
| Any smoking history – no. (%) | 639 (33.0) | 1,710 (33.5) |
| Laboratory values^†^ |  |  |
| Hemoglobin – median [IQR], g/dL | 13.7 [13.0, 14.6] | 13.8 [12.9, 14.6] |
| Platelet count – median [IQR], (10^9^ cells/L) | 236 [199, 281] | 233 [198, 275] |
| White blood cells – median [IQR], (10^9^ cells/L) | 6.8 [5.7, 8.6] | 6.7 [5.5, 8.4] |
| Mean corpuscular volume – median [IQR], fL | 90.8 [87.7, 94.0] | 90.8 [87.6, 94.0] |
| Red cell distribution width – median [IQR], % | 13.6 [13.0, 14.4] | 13.5 [12.9, 14.2] |
| Follow-up – median [IQR], year^‡^ | 2.3 [1.6, 2.9] | 2.3 [1.3, 2.9] |
| Type of incident cytopenia^§^ |  |  |
| Anemia – no. (%) | 237 (12.2) | 499 (9.8) |
| Thrombocytopenia – no. (%) | 52 (2.7) | 108 (2.1) |
| Leukopenia – no. (%) | 24 (1.24) | 61 (1.2) |
| Incident cytopenia – no. (%) | 284 (14.7) | 613 (12.0) |
| Incidence of cytopenia – per 1,000 person-years | 67 | 56 |
| Death – no. (%) | 37 (1.9) | 104 (2.0) |

IQR: interquartile range.

^*^ Cases and controls were matched 3:1 on age ± 3 years, sex, any smoking history.

^†^ Hematologic measurements were obtained from complete blood count obtained nearest to time of sequencing.

^‡^ Follow-up time is the number of years from sequencing to death or last follow-up in each cohort, whichever is earliest.

^§^ Cytopenia definitions: anemia (hemoglobin < 12.0 g/dL for females or 13.0 g/dL for males), thrombocytopenia (platelet count < 150,000 cells/μL), leukopenia (white blood cell count < 3,700 cells/μL). Cytopenias were only counted if there were two consecutive observations of a cytopenia in a single lineage at least 120 days apart without an intervening normal measurement.

**Table S2. Characteristics of cases and controls in Vanderbilt’s BioVU biorepository.^*^**

| Characteristic | **Cases**  **(N=1,622)** | **Controls**  **(N=4,314)** |
| --- | --- | --- |
| Age – median [IQR], year | 65.2 [55.6, 73.0] | 63.8 [54.6, 72.0] |
| Female – no. (%) | 915 (56.4) | 2,437 (56.5) |
| Any smoking history – no. (%) | 456 (28.1) | 1,209 (28.0) |
| Laboratory values^†^ |  |  |
| Hemoglobin – median [IQR], g/dL | 13.8 [13.0, 14.7] | 13.8 [12.9, 14.8] |
| Platelet count – median [IQR], (10^9^ cells/L) | 246 [202, 298] | 241 [202, 288] |
| White blood cells – median [IQR], (10^9^ cells/L) | 7.2 [5.9, 9.1] | 7.1 [5.8, 8.8] |
| Mean corpuscular volume – median [IQR], fL | 91.0 [88.0, 94.0] | 91.0 [88.0, 94.0] |
| Red cell distribution width – median [IQR], % | 13.6 [13.0, 14.4] | 13.4 [12.9, 14.2] |
| Follow-up – median [IQR], year^‡^ | 3.3 [1.1, 7.2] | 3.5 [1.2, 7.8] |
| Type of incident cytopenia^§^ |  |  |
| Anemia – no. (%) | 376 (23.2) | 883 (20.5) |
| Thrombocytopenia – no. (%) | 84 (5.2) | 205 (4.8) |
| Leukopenia – no. (%) | 41 (2.5) | 81 (1.9) |
| Incident cytopenia – no. (%) | 432 (26.6) | 1,014 (23.5) |
| Incidence of cytopenia – per 1,000 person-years | 58 | 48 |
| Death – no. (%) | 185 (11.4) | 407 (9.4) |

IQR: interquartile range.

^*^ Cases and controls were matched 3:1 on age ± 3 years, sex, any smoking history.

^†^ Hematologic measurements were obtained from complete blood count obtained nearest to time of sequencing.

^‡^ Follow-up time is the number of years from sequencing to death or last follow-up in each cohort, whichever is earliest.

^§^ Cytopenia definitions: anemia (hemoglobin < 12.0 g/dL for females or 13.0 g/dL for males), thrombocytopenia (platelet count < 150,000 cells/μL), leukopenia (white blood cell count < 3,700 cells/μL). Cytopenias were only counted if there were two consecutive observations of a cytopenia in a single lineage at least 120 days apart without an intervening normal measurement.

**Table S3. Characteristics of cases and controls in UK Biobank.^*^**

| Characteristic | **Cases**  **(N=5,815)** | **Controls**  **(N=15,326)** |
| --- | --- | --- |
| Age – median [IQR], year | 62.4 [57.5, 66.2] | 62.3 [57.0, 65.9] |
| Female – no. (%) | 3,198 (55.0) | 8,478 (55.3) |
| Any smoking history – no. (%) | 3,717 (63.9) | 9,720 (63.4) |
| Laboratory values^†^ |  |  |
| Hemoglobin – median [IQR], g/dL | 14.1 [13.4, 15.0] | 14.10 [13.3, 14.9] |
| Platelet count – median [IQR], (10^9^ cells/L) | 250 [216, 290] | 249 [214, 289] |
| White blood cells – median [IQR], (10^9^ cells/L) | 6.8 [5.7, 8.0] | 6.6 [5.6, 7.8] |
| Mean corpuscular volume – median [IQR], fL | 91.1 [88.5, 93.8] | 91.1 [88.6, 93.8] |
| Red cell distribution width – median [IQR], % | 13.4 [13.0, 13.9] | 13.4 [12.9, 13.9] |
| Follow-up – median [IQR], year^‡^ | 5.9 [4.7, 6.7] | 5.9 [4.8, 6.7] |
| Type of incident cytopenia^§^ |  |  |
| Anemia – no. (%) | 400 (6.9) | 917 (6.0) |
| Thrombocytopenia – no. (%) | 124 (2.1) | 274 (1.8) |
| Leukopenia – no. (%) | 71 (1.2) | 129 (0.8) |
| Incident cytopenia – no. (%) | 553 (9.5) | 1,255 (8.2) |
| Incidence of cytopenia – per 1,000 person-years | 17 | 15 |
| Death – no. (%) | 823 (14.2) | 1,693 (11.1) |

IQR: interquartile range.

^*^ Cases and controls were matched 3:1 on age ± 3 years, sex, any smoking history.

^†^ Hematologic measurements were obtained from complete blood count obtained nearest to time of sequencing.

^‡^ Follow-up time is the number of years from sequencing to death or last follow-up in each cohort, whichever is earliest.

^§^ Cytopenia definitions: anemia (hemoglobin < 12.0 g/dL for females or 13.0 g/dL for males), thrombocytopenia (platelet count < 150,000 cells/μL), leukopenia (white blood cell count < 3,700 cells/μL). Cytopenias were only counted if there were two consecutive observations of a cytopenia in a single lineage at least 120 days apart without an intervening normal measurement.

**Table S4. ICD-9 and ICD-10 codes used to define myeloid neoplasms**

| **Disease** | **ICD-9** | **ICD-10** |
| --- | --- | --- |
| Acute myeloid leukemia | 205.0, 205.00, 205.01, 205.02 | C92.0, C92.00, C92.01, C92.02, C92.4, C92.40, C92.41, C92.42, C92.5, C92.50, C92.51, C92.52, C92.6, C92.60, C92.61, C92.62, C92.A, C92.A0, C92.A1, C92.A2 |
| Essential thrombocythemia | 238.71 | D47.3 |
| Myelodysplastic syndrome | 238.72, 238.73, 238.74, 238.75 | D46, D46.0, D46.1, D46.2, D46.20, D46.21, D46.22, D46.4, D46.9, D46.A, D46.B, D46.C, D46.Z |
| Myelofibrosis | 289.83 | D47.4, D75.81 |
| Polycythemia vera | 238.4 | D45 |

ICD-9, ICD-10: International Classification of Diseases, Ninth and Tenth Revision.

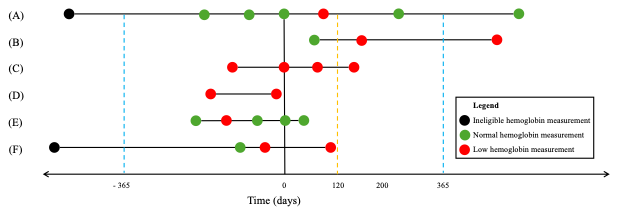

**Figure S1. Multi-timepoint complete blood count eligibility and cytopenia criteria.** Time-series plot depicting six scenarios that demonstrate criteria for study eligibility and persistent cytopenia. Time of sequencing is denoted as zero on the x-axis. The blue dashed lines represent one year prior to and after sequencing. The gold dashed line represents 120 days after sequencing. Each circle represents a hemoglobin measurement. To be eligible for the study, participants must have multi-timepoint complete blood count (CBC) data, no laboratory evidence of persistent cytopenia before sequencing, and no diagnosis of myeloid neoplasm before sequencing. Multi-timepoint CBC was defined as at least three CBC measurements, including one within a year of sequencing and two on or after the date of sequencing. The final CBC measurement had to occur at least 120 days after sequencing or the first CBC measurement, whichever came later. CBC measurements occurring greater than one year before sequencing were not included in analysis. Persistent cytopenia was defined as two consecutive observations of a cytopenia in a single lineage at least 120 days apart without an intervening normal measurement. (A) Participant with six eligible and one ineligible CBC measurement who met criteria for multi-timepoint CBC and did not have evidence of an incident, persistent cytopenia. (B) Participant with three eligible CBC measurements who met criteria for multi-timepoint CBC and an incident, persistent cytopenia. (C) Participant with four eligible CBC measurements who met criteria for multi-timepoint CBC and a prevalent, persistent cytopenia. (D) Participant with two eligible CBC measurements who did not meet criteria for multi-timepoint CBC, but did meet criteria for a prevalent, persistent cytopenia. (E) Participant with five eligible CBC measurements who did not meet criteria for multi-timepoint CBC nor persistent cytopenia. (F) Participant with three eligible and one ineligible CBC measurement who did not meet criteria for multi-timepoint CBC, but did meet criteria for a prevalent, persistent cytopenia. The participants labeled (A) and (B) would be included in the case-control study. Participant (C) would be excluded for cytopenia prior to enrollment. Participants (D), (E), and (F) would be excluded for insufficient CBC measurement data.

**Figure S2.** **Flow diagram showing selection of cases and controls across cohorts.** Abbreviations: AML = acute myeloid leukemia; MDS = myelodysplastic syndrome; MF = myelofibrosis; CBC = complete blood count; CHIP = clonal hematopoiesis of indeterminate potential. Participants were screened from (A) All of Us Research Program (N=243,609), (B) Vanderbilt’s BioVU biorepository (N=107,607), and (C) UK Biobank (N=454,033). Participants were excluded for prior AML, MDS or MF diagnoses, insufficient CBC data, or cytopenia at enrollment. Cases and controls were matched 1:3 on age ± 3 years, gender, and any history of smoking within their respective cohort. The same control was able to be matched to multiple cases.

**Figure S3. Risk of incident cytopenia by genotype across cohorts.** Abbreviations: CHIP = clonal hematopoiesis of indeterminate potential. Univariate Cox regression analyses for incident cytopenia by specific CHIP genotypes at time of enrollment for participants with CHIP and controls without CHIP serving as the reference group in (A) All of Us Research Program, (B) Vanderbilt’s BioVU biorepository, and (C) UK Biobank.

**Figure S4.** **Risk of incident cytopenia by baseline characteristic across cohorts.** Abbreviations: VAF = variant allele fraction; CHRS = clonal hematopoiesis risk score; MCV = mean corpuscular volume (femtoliters); RDW = red cell distribution width (%); CHIP = clonal hematopoiesis of indeterminate potential. Univariate Cox regression analyses for incident cytopenia by baseline characteristic at time of enrollment for participants with CHIP without adjustment. The variable high-risk genes (CHRS) indicates a participant had at least one mutation in the following genes: *SRSF2*, *SF3B1*, *ZRSR2*, *IDH1*, *IDH2*, *FLT3*, *RUNX1*, or *JAK2*. The variable high-risk genes indicates a participant had at least one mutation in the following genes: *TP53*, *PPM1D*, *SF3B1*, *SRSF2*, *U2AF1*, *ZRSR2*, *IDH1*, or *IDH2*.

**Figure S5. Risk of incident cytopenia stratified by participant risk factor profiles across cohorts.** Cumulative incidence of cytopenia in participants with clonal hematopoiesis of indeterminate potential (CHIP) in (A) All of Us Research Program, (B) Vanderbilt’s BioVU biorepository, and (C) UK Biobank stratified by the number of high-risk features at time of enrollment. High risk features were defined as age ≥ 65 years, male gender, ≥ 2 CHIP mutations, mean corpuscular volume ≥ 100 femtoliters, red cell distribution width ≥ 15%, or the presence of at least one high-risk CHIP mutation (*SRSF2, SF3B1, ZRSR2, IDH1, IDH2, FLT3, RUNX1,* or *JAK2*).
